# Parental lifespan and polygenic risk score of longevity are associated with white matter hyperintensities

**DOI:** 10.1101/2021.02.02.21251026

**Authors:** Chao Dong, Anbupalam Thalamuthu, Jiyang Jiang, Karen A. Mather, Henry Brodaty, Perminder S. Sachdev, Wei Wen

**Author notes:** Corresponding author: Chao Dong, Centre for Healthy Brain Aging, CHeBA, School of Psychiatry, UNSW Medicine, Kensington, 2052, Sydney, Australia.

## Abstract

Human longevity is moderately heritable and is hence influenced by both genetic and environmental factors. However, there remains considerable uncertainty regarding its relationship with brain ageing. In this study, we investigated the associations of parental lifespan (parental age at death) and polygenic risk score for longevity (longevity-PRS) with structural magnetic resource imaging (MRI) brain metrics considered to reflect the brain ageing process. We used a discovery sample (N = 19136) from the UK Biobank and a replication sample (N =809) from the Sydney Memory and Ageing Study and the Older Australian Twins Study. We found lower cerebral white matter hyperintensity (WMH) volumes to be significantly associated with longer parental lifespan in the discovery and replication samples and higher longevity-PRS in the discovery sample and a similar trend in the replication sample. The association of longevity-PRS with WMH remained significant after removing the influence of the *apolipoprotein E* locus. Additionally, the effects of longevity-PRS on the association were more prominent in males, especially in the older-male group. Our findings suggest that human longevity-related genes may have an influence on WMH burden, suggesting WMH volume may be a biomarker for longevity and an ageing endophenotype.

## 1. Introduction

Human longevity has increased greatly over the past centuries (Giuliani et al., 2018). Increased life expectancy has also been accompanied by an increase in age-related neurological disease burden (Feigin et al., 2017), with stroke and dementia reported in the top five primary causes of death (World Health Organization, 2018). Many age-related diseases and human longevity may share the same mechanism as brain ageing (Franceschi et al., 2020; Lockhart and DeCarli, 2014; Melzer et al., 2020; Revelas et al., 2018; Timmers et al., 2020), which are commonly characterised by grey matter (GM) atrophy and white matter hyperintensities (WMH). Therefore, the exploration of the relationships between human lifespan, longevity-related genes and ageing-related brain measures may reveal new insights.

Observational studies have shown that individuals with long-lived parents have less cognitive impairment (Andersen et al., 2019; Dutta et al., 2014), lower risk of Alzheimer’s disease (Lipton et al., 2010), healthier brain ageing processes (Murabito et al., 2014), and better-preserved brain structure (Tian et al., 2020). Furthermore, it has been shown that age-related brain changes occur over the adult lifespan, with GM and white matter (WM) atrophy (Driscoll et al., 2009; Fjell et al., 2009; Pfefferbaum et al., 2013; Raz et al., 2010; Storsve et al., 2014) and the accumulation of WMH (Wen and Sachdev, 2004; Wen et al., 2009), which are also features of neurodegeneration and cerebrovascular burden. Mounting evidence shows that periventricular WMH (PVWMH) and deep WMH (DWMH) which are two subclassifications of WMH have different neuropathology, risk factors, associations with cognition (Sachdev et al., 2005), and different genetic underpinnings (Armstrong et al., 2020). In addition, GM and WMH volumes also show sex differences (Alqarni et al., 2021; Liu et al., 2020; Lotze et al., 2019; Sachdev et al., 2009b). Therefore, the empirical evidence indicates that GM and WMH volumes are well-established brain neuroimaging metrics associated with brain ageing. Although the relationship between parental longevity and the ageing brain has been explored (Tian et al., 2020), the associations between these ageing-related metrics and longevity-related genes, and the age and sex differences in these associations remain unclear.

Recent genome-wide association studies (GWAS) have identified putative longevity candidate loci (Deelen et al., 2019; Timmers et al., 2019; Timmers et al., 2020). Polygenic risk scores (PRS) are one way to summarise results from genetic association studies for prediction of complex traits, such as longevity. Polygenic risk score for longevity (longevity-PRS) was recently shown to be associated with cognitively healthy ageing and prolonged survival (Tesi et al., 2020), and a previous study applied PRS of different clinical biomarkers to predict lifespan (Sakaue et al., 2020). The association of longevity genes with ageing-related brain metrics has not previously been reported.

In this study, we aimed to: (1) investigate the associations between parental lifespan (parental age at death) and participants’ brain macrostructure volumes, i.e. brain soft tissue volume, cortical volume, total grey matter volume, hippocampus volume, total cerebral WM volume and WMH volume (whole WMH, periventricular WMH and deep WMH); (2) investigate the associations between longevity-PRS and these brain metrics, and (3) explore age and sex differences in these associations. We performed the analysis using 19136 individuals in the UK Biobank and replicated the results in 809 individuals in the combined Sydney Memory and Ageing Study (MAS) and Older Australian Twins Study (OATS) sample.

## 2. Methods

### 2.1 Participants

In the current study, we used the UK Biobank as our discovery sample. We then combined MAS and OATS as a replication sample.

#### UK Biobank

The UK Biobank is a large-scale dataset containing both genetic and cross-modality neuroimaging data. It is a population-based study which consists of over 500,000 participants aged between 40–70 years at study entry (Sudlow et al., 2015). Around four years after initial recruitment, a subset of participants underwent MRI. Written consent was acquired from all participants and ethics approval was provided by the National Health Service National Research Ethics Service (11/NW/0382). All data presented in this analysis were collected on two scanners. After excluding participants whose MRI data or genetic data or parental lifespan data were missing, 19136 participants were included in the final analysis (including parental lifespan and longevity-PRS analysis). All data and materials are available via UK Biobank (http://www.ukbiobank.ac.uk). Our study was approved in October 2018 (Application number: 37103),

#### Sydney Memory and Ageing Study (Sydney MAS)

Sydney MAS is a cohort of 1037 community dwelling adults aged 70–90 years (Sachdev et al., 2010). Five hundred and thirty-nine of the 1037 participants received MRI scans at baseline (Wave 1). In the two-year (Wave 2) follow-up, MRI data were acquired for 415. For the present study, participants from Wave 1 and 2 were used (we chose Wave 2 scans when there were overlapping, as Wave 2 scans were considered to be of superior quality). Ethics approval was obtained from the Human Research Ethics Committees of the University of New South Wales and the South Eastern Sydney Local Health District. More information about the derivation of the final analytic sample can be found in Figure S1 (Supplementary material) and Table 1.

**Table 1.** Sample characteristics.

|  | UK Biobank<br>(discovery sample) |  | MAS & OATS<br>(replication sample) |  |
| --- | --- | --- | --- | --- |
|  | Parental lifespan<br>N=16796 | Longevity-PRS<br>N=18606 | Parental lifespan<br>N=768 | Longevity -PRS<br>N=773 |
| **Brain volumes** | N=16639 | N=18435 | N=754 | N=760 |
| Age, years | 63.6 (7.0) | 62.5 (7.5) | 78.3 (5.2) | 78.3 (5.3) |
| Male | 7872 (47.3%) | 8654 (46.9%) | 330 (43.8%) | 331 (43.6%) |
| Cortex, mm<sup>3</sup> | 458011.8 (41840.9) | 459427.5 (41963.9) | 434473.5 (54953.3) | 432850.8 (55639.5) |
| Total Gray matter, mm<sup>3</sup> | 628438.5 (53411.1) | 630281.2 (53590.2) | 582265.5 (63619.2) | 580648.5 (64307.2) |
| Brain soft tissue, mm<sup>3</sup> | 1131294 (105213.1) | 1134623 (105525.3) | 1015695 (109848.5) | 1013977 (111225) |
| Hippocampus, mm<sup>3</sup> | 8170.4 (778.5) | 8202.1 (781.1) | 7129.8 (822.0) | 7117.6 (832.5) |
| Cerebral WM, mm<sup>3</sup> | 473416.2 (55123.5) | 474791.2 (55213.2) | 406129.7 (53657.1) | 406065.8 (54453.7) |
| **WMH volumes** | N=14991 | N=16583 | N=725 | N=728 |
| Age, years | 63.2 (7.0) | 62.1 (7.4) | 78.3 (5.2) | 78.2 (5.2) |
| Male | 7098 (47.3 %) | 7788 (47.0%) | 321 (44.3%) | 321 (44.1%) |
| Whole WMH, mm<sup>3</sup> | 1875.6 (1669.0) | 1764.1 (1577.1) | 15836.8 (12833.6) | 15818 (12808.9) |
| PVWMH, mm<sup>3</sup> | 1427.3 (1308.3) | 1344.1 (1234.8) | 11221.2 (8587.9) | 11179.6 (8535.2) |
| DWMH, mm<sup>3</sup> | 425.3 (483.6) | 397.9 (457.9) | 4372.8 (5047.4) | 4396.2 (5088.9) |

Notes. Mean (standard deviation) or N (%)
Brain soft tissue (labelled as BrainSegVolNotVent in FreeSurfer); WM: white matter; WMH: white matter hyperintensity; PVWMH: periventricular WMH; DWMH: deep WMH. MAS: Sydney Memory and Ageing Study; OATS: Older Australian Twins Study.

#### Older Australian Twins Study (OATS)

OATS is a study of community-dwelling twins aged 65 and above. The recruitment of participants occurred in three Eastern states of Australia: New South Wales, Victoria, and Queensland. This study was approved by the ethics committees of the Australian Twin Registry, University of New South Wales, University of Melbourne, Queensland Institute of Medical Research and the South Eastern Sydney & Illawarra Area Health Service. Informed consent was obtained from all subjects and the methods were carried out in accordance with the relevant guidelines. At baseline, 623 individuals had participated and 400 of them had brain MRI data. Data was collected every two years. Methodology of OATS has previously been described in detail (Sachdev et al., 2009a; Sachdev et al., 2013). Waves 1-Wave 4 data were used in the current study, and we selected only one individual from each family. In addition, as there were more female participants than males, we selected a male participant for inclusion in this study when both female and male participants were available from the same family. More information about the derivation of final analytic sample can be found in Figure S1 (Supplementary material) and Table 1.

### 2.2 Neuroimaging data and pre-processing

#### UKB

We used T1-weighted and FLAIR scans and the key parameters for MRI imaging were: (i) T1-weighted MRI: TI = 880 ms, TR = 2,000 ms, resolution = 1.0 × 1.0 × 1.0 mm, matrix size = 208 × 256×256. (ii) T2-weighted FLAIR: TI = 1800 ms, TR = 5,000 ms, resolution = 1.05 × 1.0 × 1.0 mm, matrix size = 192 × 256×256. T1-weighted scans for 19363 participants were downloaded in November 2018.

#### Sydney MAS

We used T1-weighted and FLAIR scans and the key parameters for MRI imaging were: (i) T1-weighted MRI: TR = 6.39 ms, TE = 2.9 ms, flip angle = 8°, matrix size = 256×256, FOV (field of view) = 256×256×190, and slice thickness = 1 mm with no gap in between, yielding 1×1×1 mm^3^ isotropic voxels; (ii) T2-weighted FLAIR: TR = 10000 ms, TE =110 ms, TI = 2800 ms, matrix size = 512×512, slice thickness = 3.5 mm without gap, and in plane resolution = 0.488×0.488 mm.

#### OATS

The neuroimaging data were obtained on the five scanners because of the multi-site nature of this study. The detailed scanning parameters are presented in Table S1 (Supplementary material).

Using T1-weighted images from UK Biobank, MAS and OATS, the reconstruction of pial surface, grey matter (GM)/ white matter (WM) boundary and WM/subcortical GM boundary was automatically performed by using FreeServer ver.7.1.0 (Fischl, 2012), to quantify volumes of whole brain soft tissue (labelled as BrainSegVolNotVent in FreeSurfer), total GM, total WM, cortex, and hippocampi. The segmentation and quantification of WMH in all three cohorts were conducted using UBO Detector (Jiang et al., 2018). We applied the pipeline with default settings for MAS and OATS. Visual inspection of the results showed good segmentation accuracy. However, we found the default UBO Detector did not perform well in UK Biobank mainly due to the presence of subtle WMH in this younger cohort compared to MAS and OATS. Therefore, we modified our UBO Detector to extract WMH in the individual FLAIR space. All atlases and masks in the standard space were transformed to individual space by applying the reversed flow map. Since the WMH extraction in MAS and OATS was done in a standard space, intracranial volume (ICV) was not accounted for in statistical analyses. The WMH measures in UK Biobank, on the other hand, were segmented in the individual space and biased by inter-subject differences in brain sizes. Therefore, ICV was adjusted for in all regression models when testing WMH in UK Biobank. In UK Biobank, MAS and OATS, periventricular WMH (PVWMH) was defined as WMH voxels within 12 mm of distance from the nearest voxel in the lateral ventricle, and the rest WMH voxels were classified as deep WMH (DWMH) (Jiang et al., 2018). This distance threshold is comparable to those commonly used in automated PVWMH/DWMH separation (Griffanti et al., 2018).

### 2.3 Parental lifespan

In the present study, parental age at death was determined through self-report by participants, and parental lifespan was defined as parental age at death. Specifically, if parental lifespan data was missing for one parent, the other parent’s lifespan was used as the parental lifespan. Otherwise, parental lifespan was calculated as the mean value of the father and mother’s lifespan. Participants were excluded if both father and mother’s attained age (dead or alive) was less than 40 years old, as death in such cases was likely to have been accidental.

As supplementary analyses, we also did paternal analysis (father’s age only) and maternal analysis (mother’s age only) to explore their associations with brain metrics separately.

### 2.4 Genotyping and Imputation

#### UK Biobank

Genotyped and imputed data for approximately 500,000 samples were available. The details of genotyping and imputation in UK biobank samples can be found elsewhere (Bycroft et al., 2018; McCarthy et al., 2016). Briefly, the genotyping was performed using the Bileve or UK Biobank axiom arrays. Imputation was undertaken using the Haplotype Reference Consortium (HRC), 1000 genome and UK Biobank reference panels. For PRS calculation only those SNPs which remained after quality control (QC) filtering (MAF > 0.1%, imputation information score > 0.6) were used. Also, we have used only those samples with reported British ancestry and removed any samples with high genotype missing rate and relatedness.

#### Sydney MAS and OATS

Genotyping in MAS and OATS samples were performed using Affymetrix SNP 6.0. or the Illumina Ominexpress SNP arrays, respectively. Genotyped SNPs were QCed using standard procedures (>95% call rate; HWE p-value > 10^−6^; MAF > 0.01; non-strand ambiguous SNPs). Ethnic outliers were identified and removed using principal components analysis. Whole genome imputation was performed using the Michigan imputation server (https://imputationserver.sph.umich.edu) based on the HRC r1.1 2016 (GRch 37/hg19). Imputed SNPs were QCed as for the UK Biobank.

### 2.5 Longevity polygenic risk scoring

Longevity GWAS (Deelen et al., 2019) summary statistics were downloaded from the GWAS catalogue (https://www.ebi.ac.uk/gwas/downloads/summary-statistics). The longevity PRS was calculated using the program PRS-CS (Ge et al., 2019), which uses the Bayesian regression framework with continuous shrinkage priors to obtain posterior effect sizes of the summary statistics. It produces a pruned set of effect sizes using linkage disequilibrium (LD) among SNPs based on a reference panel. For the present study we have used the precompiled LD pattern of the 1000 Genome European reference panel with the default parameters of PRS-CS. The pruned set of effect sizes from the whole genome (without the need to use thresholds based on the GWAS p-values) was used to generate PRS scores using PLINK software (Chang et al., 2015). Additionally, as the *apolipoprotein E* gene (*APOE*) locus is known to be significantly associated with neurodegenerative disease and longevity (Abondio et al., 2019; Deelen et al., 2019; Franceschi et al., 2020; Revelas et al., 2018), we also examined whether the observed effects were purely due to the *APOE* locus by excluding all the SNPs around the *APOE* locus (GRch 37; 19: 45116911:46318605).

### 2.6 Parental lifespan and brain metrics samples

In the UKB, a total of 20960 had parents’ attained age at death documented. After merging with MRI brain metrics data and then removing outliers (> 3σ), 16796 participants were included in the final parental lifespan analysis. Similarly, in replication sample, there were 1555 with parents’ attained age documented and 768 were included in the final analysis.

### 2.7 Longevity-PRS and brain metrics samples

In the discovery sample, 23200 participants had genetic data available. There were 18606 participants in the final longevity-PRS analysis after merging with brain metrics data and then removing outliers. In the replication sample, 1464 participants had genetic data and 773 were included in the final longevity-PRS analysis. More information about the derivation of the final analytic sample is shown in Figure S1 (Supplementary material) and Table 1.

### 2.8 Statistical analysis

All statistical analyses were performed using R version 4.0.0. Association tests were performed using linear regression models. Longevity-PRS was z-transformed and set as the independent variable, and brain metrics as dependent variables. For parental lifespan analysis, covariates included age, age^2^, sex, age × sex (i.e. age by sex interaction), age^2^ × sex (i.e. age^2^ by sex interaction), ICV, scanner. Apart from parental lifespan, we also undertook maternal and paternal lifespan analyses separately, controlling for the same covariates. For the longevity-PRS analysis, we controlled for all the above covariates and 10 genetic principal components (PCs). Additionally, we controlled for ICV in all regression models when testing WMH in UK Biobank as WMH measures in the UK Biobank were segmented in the individual space and biased by inter-subject differences in brain sizes. Regression coefficients (β) are reported as the effect sizes of the parental lifespan or longevity-PRS. False Discovery Rate (FDR) values were obtained by using the Benjamini-Hochberg (1995) procedure and the significant threshold was set as FDR ⩽0.05.

In addition, the discovery sample was divided into PRS quartiles to investigate differences between the PRS groups with the highest and lowest PRS, using the highest PRS group as the reference. As the age range and the number of participants were large in the UK Biobank, we also explored age and sex effects by splitting these participants into two age groups using the median age of 63.

## 3. Results

Figure 1 provides an overview of the datasets. Two main analyses were undertaken in the present study, which examined the associations between brain metrics and (i) parental lifespan and (ii) longevity-PRS in both the discovery and replication samples. More detailed information on the selection of samples is described in Figure S1 (Supplementary). Table 1 summarises the characteristics of the samples. The discovery sample had a mean age of ∼63 years compared to 78 years in the replication cohorts.

**Figure 1.**
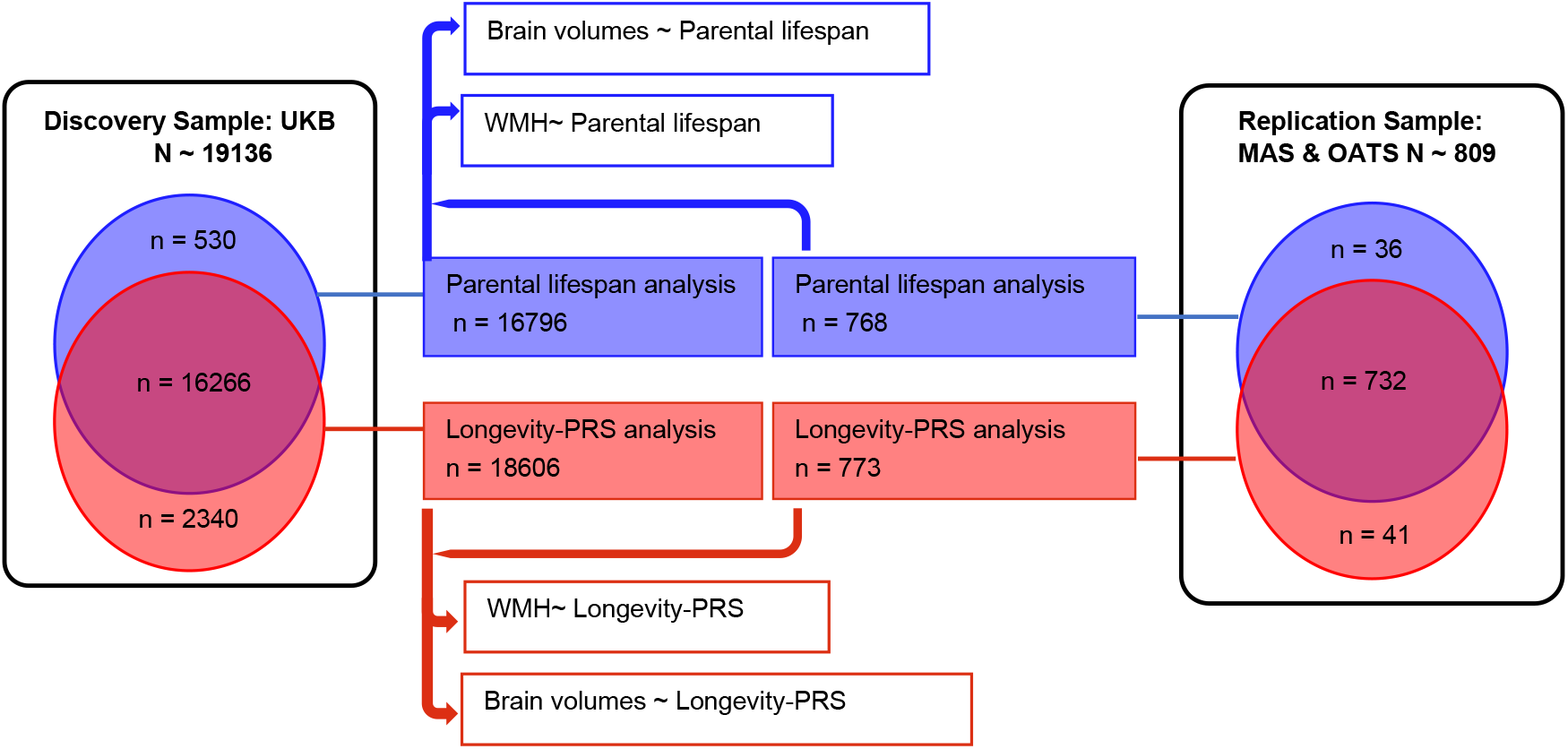
Overview of the study. The left and right rectangles represent discovery and replication samples. Two main analyses were undertaken examining brain metrics with (i) parental lifespan analysis (blue circle) and (ii) longevity-PRS analysis (orange circle). The overlap of samples (red) between the two analyses is shown separately for the discovery and replication samples. All numbers represent the number of participants in each analysis (see Table 1 for more detailed information). UKB: UK Biobank; MAS: Sydney Memory and Ageing Study; OATS: Older Australian Twins Study.

### 3.1 Associations between parental lifespan and brain metrics

In the discovery sample (Figure 2. A), parental lifespan was significantly associated with cortical volume (β=0.0105, P_adj_ =0.0174), total grey matter volume (β=0.0124, P_adj_ =0.0030), and three WMH volumes: whole WMH (β=-0.0323, P_adj_ =0.0002), PVWMH (β=-0.0305, P_adj_ =0.0003), and DWMH (β=-0.0273, P_adj_ =0.0015), while brain soft tissue volume, total cerebral white matter volume, and hippocampal volume did not show significant associations (P>0.05) (Table 2). In addition, we found that the association of WMH with paternal lifespan appeared to be stronger compared to maternal lifespan (Supplementary Table S2).

**Table 2.** Associations between brain metrics volumes and parental lifespan.

| Phenotypes | UK Biobank<br>(discovery sample) |  |  |  | MAS & OATS<br>(replication sample) |  |  |  |
| --- | --- | --- | --- | --- | --- | --- | --- | --- |
| | $\beta$ | SE | P | P<sub>adj</sub> | $\beta$ | SE | P | P<sub>adj</sub> |
| Cortex | **0.0105** | **0.0041** | **0.0109\*** | **0.0174\*** | 0.0229 | 0.0271 | 0.3993 | 0.6277 |
| Total Gray matter | **0.0124** | **0.0039** | **0.0015\*** | **0.0030\*** | 0.0150 | 0.0251 | 0.5493 | 0.6277 |
| Brain soft tissue | 0.0049 | 0.0033 | 0.1436 | 0.1914 | 0.0150 | 0.0221 | 0.4976 | 0.6277 |
| Hippocampus | 0.0043 | 0.0063 | 0.4994 | 0.4994 | 0.0017 | 0.0312 | 0.9573 | 0.9573 |
| Cerebral WM | -0.0039 | 0.0042 | 0.3522 | 0.4026 | 0.0171 | 0.0225 | 0.4454 | 0.6277 |
| Whole WMH | **-0.0323** | **0.0076** | **2.47E-05\*** | **0.0002\*** | **-0.0871** | **0.0311** | **0.0052\*** | **0.0208\*** |
| PVWMH | **-0.0305** | **0.0077** | **0.0001\*** | **0.0003\*** | **-0.0880** | **0.0311** | **0.0048\*** | **0.0208\*** |
| DWMH | **-0.0273** | **0.0079** | **0.0006\*** | **0.0015\*** | -0.0700 | 0.0331 | 0.0346* | 0.0924 |

Notes. Brain soft tissue (labelled as BrainSegVolNotVent in FreeSurfer); WM: white matter; WMH: white matter hyperintensity; PVWMH: periventricular WMH; DWMH: deep WMH. Asterisks represent statistical significance when using linear mixed models (FDR < 0.05). SE: standard error. P<sub>adj</sub>: False Discovery Rate. MAS: Sydney Memory and Ageing Study; OATS: Older Australian Twins Study.

**Figure 2.**
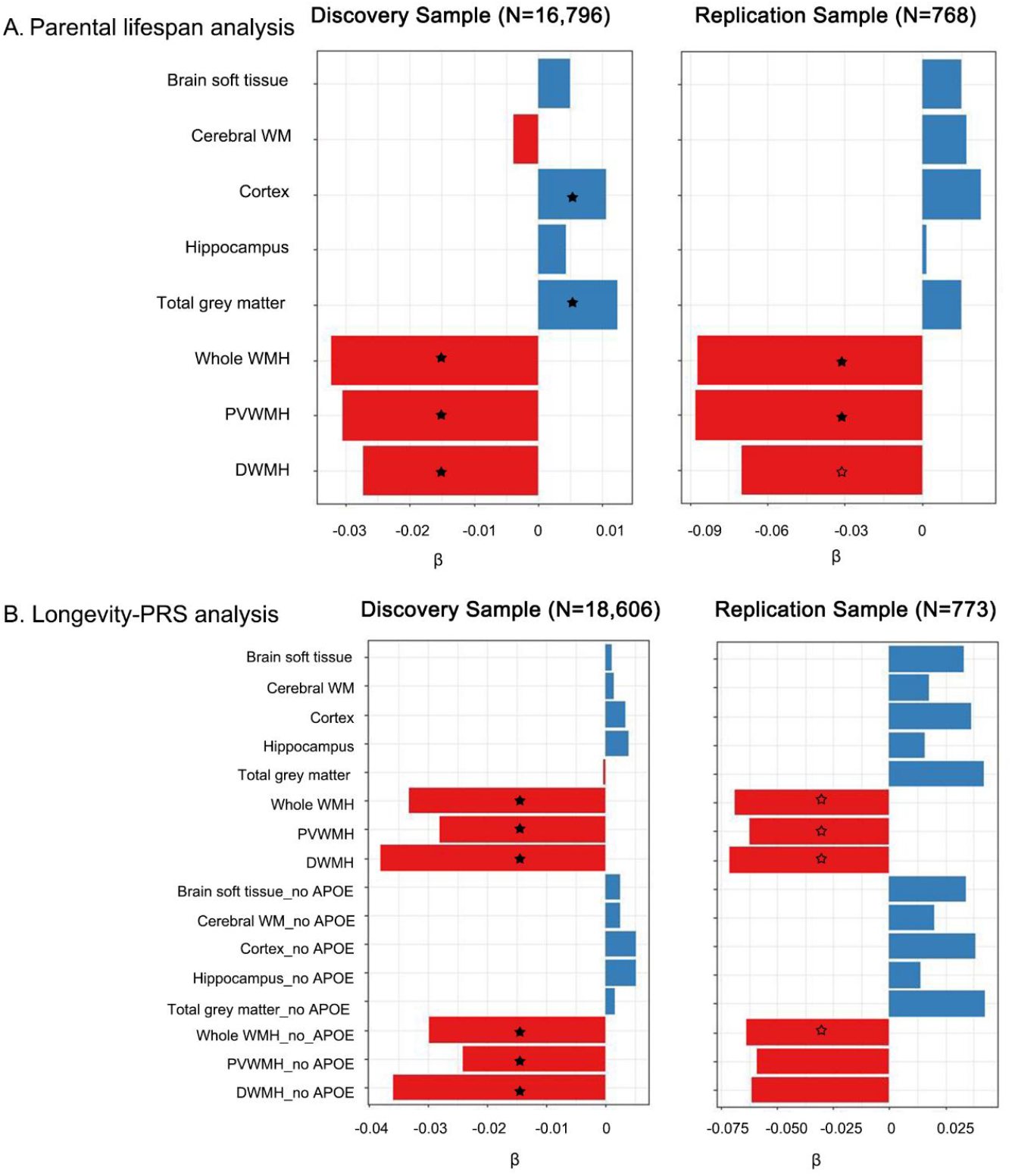
Comparisons of effect sizes for parental lifespan and lifespan-PRS with brain metrics. **A**. Parental lifespan and brain metric analyses for discovery and replication samples. **B**. Longevity-PRS and brain metric analyses for discovery and replication samples. X axes represent the effect size (β). Measures with solid asterisk were significantly associated with parental lifespan or longevity-PRS after False Discovery Rate (FDR) correction (P_adj_ <0.05), and measures with a hollow asterisk represent those associations that were significant (P <0.05) but did not survive FDR correction.

In the replication sample, parental lifespan was also associated with whole WMH (β=-0.0871, P_adj_ =0.0208), PVWMH (β=-0.0880, P_adj_ =0.0208), while parental lifespan associations with other brain metrics were not significant (P_adj_ > 0.05). Importantly, the effect sizes (β) showed similar direction in the discovery and replication samples, especially for WMH (Table 2, Figure 2. A).

### 3.2 Associations between longevity-PRS and brain imaging metrics

Table 3 and Figure 2. B summarises the main findings of the association between longevity-PRS and brain metrics volumes. In the discovery sample, we found significant associations between longevity-PRS and WMH volumes: whole WMH (β=-0.0331, P_adj_ =0.0015), PVWMH (β=-0.0279, P_adj_ =0.0079), and DWMH (β=-0.0380, P_adj_ =0.0009). There were no significant associations between longevity-PRS and other brain volumes. To determine whether the effect of longevity-PRS on these brain metrics was independent of the *APOE* locus, we tested the association of longevity-PRS excluding this locus. We found that the longevity-PRS without the *APOE* locus was also significantly associated with these WMH measures: whole WMH (β=-0.0297, P_adj_ =0.0048), PVWMH (β=-0.0241, P_adj_ =0.0235), and DWMH (β=-0.0358, P_adj_ =0.0012).

**Table 3.** Associations between brain metrics volumes and longevity-PRS.

| Phenotypes | UK Biobank<br>(discovery sample) |  |  |  |  |  |  |  | MAS & OATS<br>(replication sample) |  |  |  |  |  |  |  |
| --- | --- | --- | --- | --- | --- | --- | --- | --- | --- | --- | --- | --- | --- | --- | --- | --- |
| | $\beta$<br>(with/no *APOE*) | | SE<br>(with/no *APOE*) | | P<br>(with/no *APOE*) | | $P_{adj}$<br>(with/no *APOE*) | | $\beta$<br>(with/no *APOE*) | | SE<br>(with/no *APOE*) | | P<br>(with/no *APOE*) | | $P_{adj}$<br>(with/no *APOE*) | |
| Cortex | 0.0034 | 0.0052 | 0.0050 | 0.0050 | 0.4929 | 0.3007 | 0.8106 | 0.6872 | 0.0365 | 0.0382 | 0.0268 | 0.0273 | 0.1740 | 0.1610 | 0.2320 | 0.2320 |
| Total grey matter | -0.0002 | 0.0016 | 0.0047 | 0.0047 | 0.9623 | 0.7290 | 0.9623 | 0.8424 | 0.0422 | 0.0427 | 0.0249 | 0.0253 | 0.0910 | 0.0921 | 0.1843 | 0.1843 |
| Brain soft tissue | 0.0011 | 0.0025 | 0.0040 | 0.0040 | 0.7897 | 0.5410 | 0.8424 | 0.8106 | 0.0331 | 0.0343 | 0.0222 | 0.0226 | 0.1362 | 0.1292 | 0.2179 | 0.2179 |
| Hippocampus | 0.0040 | 0.0053 | 0.0076 | 0.0076 | 0.5961 | 0.4834 | 0.8106 | 0.8106 | 0.0160 | 0.0141 | 0.0310 | 0.0315 | 0.6061 | 0.6541 | 0.6465 | 0.6541 |
| Cerebral WM | 0.0015 | 0.0026 | 0.0050 | 0.0050 | 0.7643 | 0.6080 | 0.8424 | 0.8106 | 0.0178 | 0.0202 | 0.0226 | 0.0230 | 0.4301 | 0.3802 | 0.4915 | 0.4679 |
| Whole WMH | **-0.0331** | **-0.0297** | **0.0091** | **0.0092** | **0.0003\*** | **0.0012\*** | **0.0015\*** | **0.0048\*** | -0.0691 | -0.0637 | 0.0304 | 0.0310 | 0.0235* | 0.0401* | 0.1633 | 0.1633 |
| PVWMH | **-0.0279** | **-0.0241** | **0.0092** | **0.0092** | **0.0025\*** | **0.0088\*** | **0.0079\*** | **0.0235\*** | -0.0622 | -0.0591 | 0.0304 | 0.0309 | 0.0408* | 0.0566 | 0.1633 | 0.1766 |
| DWMH | **-0.0380** | **-0.0358** | **0.0094** | **0.0094** | **0.0001\*** | **0.0002\*** | **0.0009\*** | **0.0012\*** | -0.0714 | -0.0614 | 0.0327 | 0.0334 | 0.0294* | 0.0662 | 0.1633 | 0.1766 |

Notes. Values of $\beta$ , SE, P, $P_{adj}$ in the analysis with and no *APOE* were presented in the table. Brain soft tissue (labelled as BrainSegVolNotVent in FreeSurfer); WM: white matter; WMH: white matter hyperintensity; PVWMH: periventricular WMH, DWMH: deep WMH. Asterisks represent statistical significance when using linear mixed models ( $P < 0.05$ ). SE: standard error. $P_{adj}$ : corrected for False Discovery Rate. MAS: Sydney Memory and Ageing Study; OATS: Older Australian Twins Study.

For the PRS quartiles analysis, when compared to the highest quartile for the longevity-PRS group (reference, n= 4099), the participants with the lowest PRS (n=4147) had increased whole WMH (P_adj_ = 0.0157), PVWMH (P_adj_ = 0.0494), and DWMH (P_adj_ = 0.0056). Further, these effects on whole WMH (P_adj_ = 0.0494) and DWMH (P_adj_ = 0.0157) persisted after removing the *APOE* locus (Supplementary Table S3). This result further confirmed that participants with higher longevity-PRS had lower WMH.

In the replication sample, the associations between longevity-PRS and WMH measures showed similar significant associations in the same direction as found in discovery sample, but they did not survive multiple testing adjustment: whole WMH (β=-0.0691, P =0.0235), PVWMH (β=-0.0622, P =0.0408), and DWMH (β=-0.0714, P =0.0294). After removing the *APOE* locus, the significance of associations between longevity-PRS and WMH became weaker but showed a similar trend with whole WMH (β=-0.0637, P =0.0401), PVWMH (β=-0.0591, P =0.0566), and DWMH (β=-0.0614, P =0.0662) (Table 3). The longevity-PRS effect sizes (β) for WMH showed very similar trends across all samples (Figure 2. B).

### 3.3 Age and sex differences of longevity-PRS on WMH

In analysing the relationship between longevity-PRS and WMH, we found that the interaction between age and sex (age × sex) was significant (Supplementary Table S4). Therefore, we carried out the same analysis in males and females separately in the discovery sample, adjusting for age, age^2^, ICV, scanner and 10 genetic PCs. Significant associations between longevity-PRS and WMH were mainly found in males, which was independent of the *APOE* locus (Table 4). To further investigate sex differences of these three WMH measures in the two subgroups by age (younger group, age <63 years, n=8252; older group, age ⩾63 years, n=8331), we performed the same regression analysis separately. Significant associations were mainly found in the older-male (⩾63 years old, male) group regardless of the *APOE* locus (Table 4).

**Table 4.**
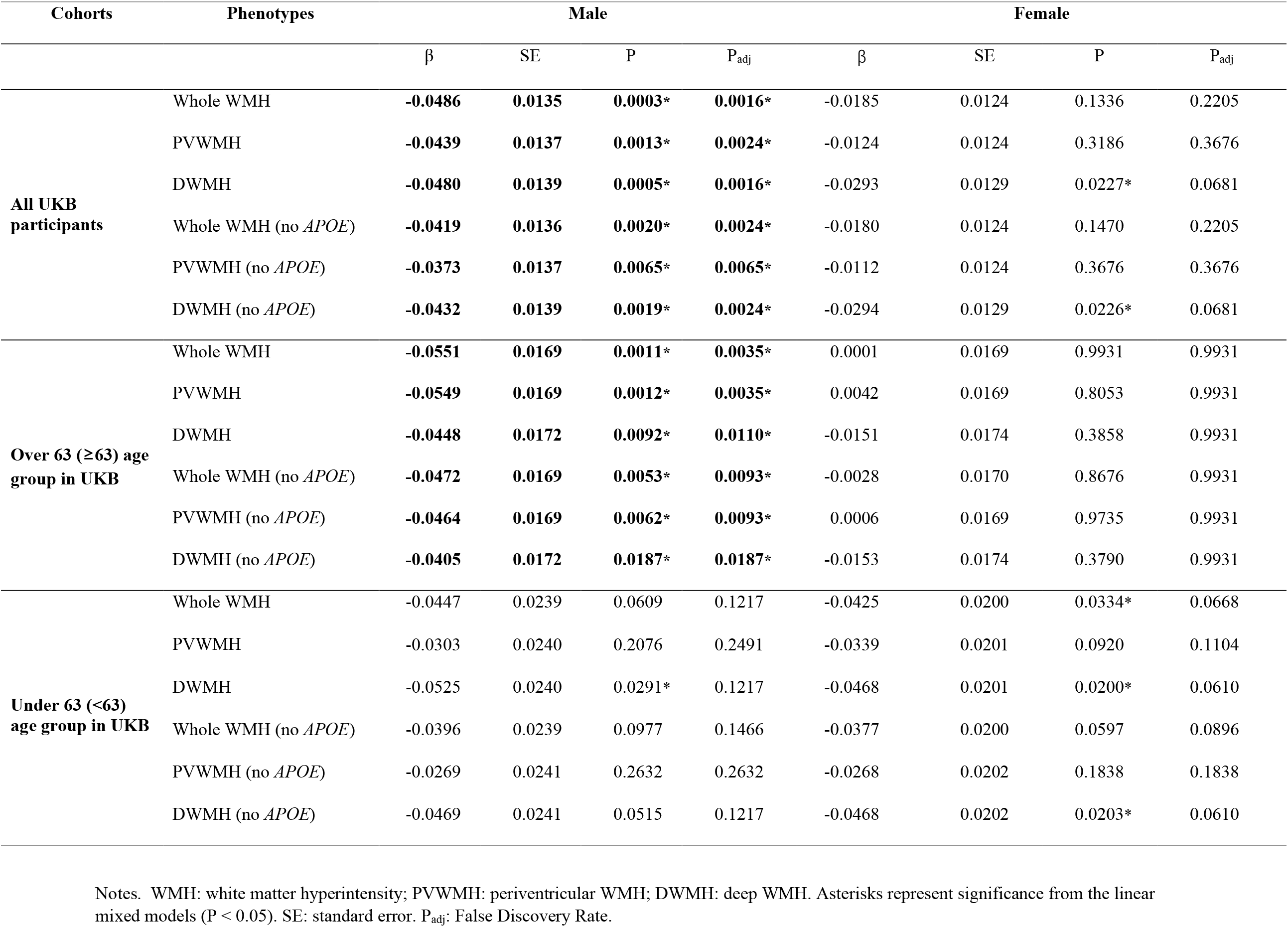
Age and sex differences in associations between WMH volumes and longevity-PRS in UK Biobank.

As age × sex was also significantly associated with other brain volumes, we explored age and sex differences on their associations with the longevity-PRS. There were no significant associations in different age-sex groups except for positive associations with cortical volume in the male group (Supplementary Table S5).

As the interaction terms between age and sex (age × sex) was not significantly associated with brain metrics in the replication sample, we did not further analyse age and sex differences of longevity-PRS effects on brain metrics.

## 4. Discussion

This study was the first to investigate associations of parental lifespan and longevity-PRS with brain imaging metrics that change with ageing. We found that parental lifespan was significantly associated with WMH volume in both the discovery and replication samples and with cortical and total grey matter volumes in the discovery sample only. Further, we found an inverse association of longevity-PRS not only with whole brain WMH, but also periventricular and deep WMH volumes separately, and these associations persisted after removing the contribution of the *APOE* locus in the large discovery sample. Similar results for the longevity-PRS were observed in the smaller replication cohort, but these did not survive multiple testing correction. In addition, sex differences were observed in the discovery cohort, with the effects of longevity-PRS on WMH being prominent in males but not females, especially in the older-male group.

These negative associations of the longevity measures with WMH are consistent with previous work (Murabito et al., 2014) demonstrated that parental longevity was associated with lower odds of extensive WMH, and exceptional familial longevity has been associated with lower susceptibility to WMH (Altmann-Schneider et al., 2013). It is noteworthy that our study examined parental lifespan after age 40 and did not specifically investigate exceptional longevity, unlike the Murabito et al study that used 85 years as the cut-off to define parental longevity. Our study therefore broadens the findings to parents who die at younger ages, showing that the relationship to brain health of offspring is not restricted to the exceptionally long-lived. These results are further suggested in the longevity-PRS analysis, which in fact is based on exceptional longevity, with long-lived cases defined as surviving to an age beyond the 90^th^ survival percentiles (Deelen et al., 2019).

Our finding of a lack of association of the longevity measures with cortical and grey matter volumes is also consistent with the aforementioned study that examined the association with parental longevity (⩾85 yrs) (Murabito et al., 2014). However, in another study also using UK Biobank data (Tian et al., 2020), parental longevity was associated with some brain structures, including hippocampal volumes, which we did not find in the present study. The Tian et al study again defined parental longevity differently using age 85 as the cut-off, unlike our examination in relation to parental lifespan beyond 40 years, which we analysed as a continuous trait. That we did not find an association of longevity-PRS with cortical, grey matter and hippocampal volumes is consistent with the Murabito et al but not the Tian et al report. The methodological differences could explain these discrepancies, particularly in the selection of samples and the use of a longevity-PRS, since the latter currently captures only a fraction of the variance in parental longevity.

Nevertheless, our analysis suggests that the association of parental age with the phenotype of WMH in the offspring may be a more robust association than that with brain volumetric measures. That WMH are associated with parental lifespan and share their genetic architecture with longevity is not surprising. It is known that WMH are highly heritable (Sachdev et al., 2016). WMH are generally considered to be neuroimaging indicators of cerebral small vessel disease, although they also have contributions from neurodegeneration, inflammation, and other possible mechanisms (Alber et al., 2019) and are regarded as an indicator of poor brain health (Wardlaw et al., 2015). The presence of WMH indicates an increased risk of stroke, dementia and death (Debette and Markus, 2010). Factors such as hypertension and diabetes are known vascular risk factors of WMH (Jorgensen et al., 2018). The disease phenotypes associated with WMH are major causes of death and thereby are linked to longevity. It is recognised that longevity-related phenotypes are genetically correlated with several disease-related phenotypes (such as high-density lipoprotein cholesterol), and our study adds WMH to this list. WMH may therefore be considered a biomarker of longevity in future studies. It must be pointed out that the magnitude of the associations is not large, suggesting that the variance explained by the shared genetics between longevity and WMH is small indeed. WMH is a complex phenotype with several determinants of which longevity-related genetics is but one. Moreover, there is some evidence of genetic and pathophysiological differences between PVWMH and DWMH (Armstrong et al., 2020), explaining different strengths in their relationship with the longevity-PRS, with DWMH showing a stronger relationship in the current study.

The longevity-PRS was derived from a meta-analysis of multiple studies (Deelen et al., 2019). Of the individual candidate genes, the only locus that appears to be significant in multiple independent meta-analyses is the *apolipoprotein E* (*APOE*) locus, with *APOE-ε4* being associated with lower odds of longevity, and *APOE-ε2* showing the opposite effect (Revelas et al., 2018). The *APOE* polymorphism is associated with cardiovascular disease and Alzheimer’s disease (Serrano-Pozo et al., 2021), with *ε4* and *ε2* having opposite effects, with the former being deleterious and the latter protective. In our study, the association of the longevity-PRS with WMH was shown to be independent of the *APOE* polymorphism, highlighting the importance of other genes and polymorphisms. These genes have been previously correlated with several disease phenotypes such as coronary artery disease, diabetes and obesity, all of which are age-related and have been linked with lifespan (Timmers et al., 2019).

In the UK Biobank sample, we found sex differences in the associations between longevity-PRS and WMH, with the longevity-PRS effects on WMH becoming stronger in males with increasing age. Previous studies have found differences in severity and heritability of WMH in men and women, with heritability also varying with age (Atwood et al., 2004; Sachdev et al., 2009b; Sachdev et al., 2016; ten Kate et al., 2018). These sex differences in WMH may be one reason why the longevity-PRS is differently associated with WMH in males and females. In addition, the PRS of extreme human longevity has been shown previously to have a larger effect on male-survival compared to female, with males depending more on an advantageous genetic background to reach extreme ages (Tesi et al., 2020). This may partly explain our finding that the effect size (β) of longevity-PRS on WMH is larger in the older-male group compared with the older-female group. In relation to the association of longevity PRS with brain volumetrics, while no significant results were found in the overall analysis, a subsidiary exploratory sex-specific analysis was performed. We found cortical volume to be positively associated with longevity-PRS (P_adj_ = 0.0066, Supplementary Table S5) in males, especially in the younger-male group (P_adj_ = 0.0214, Supplementary Table S5). We regard this finding as tentative as it was not replicated.

Our study has a number of strengths. The large and well-defined sample is a unique strength of the UK Biobank. We were able to replicate the major findings in an independent sample using the same analytical protocol for the neuroimaging parameters. We used two different strategies to examine the genetic contribution to longevity. The longevity-PRS was based on a longevity GWAS, where long-lived cases who had achieved the 90^th^ percentile in lifespans for their age cohorts, while parental lifespan used parental age of death as a continuum if the age of 40 at death had been achieved. The findings therefore suggest that WMH in offspring are related to both exceptional longevity as well as the average lifespan of parents, suggesting its robustness as a marker of longevity.

This study also has several limitations. First, both of the longevity measures (parental longevity, longevity-PRS) have shortcomings. Parental lifespan was based on self-report and is therefore prone to reporting error. The longevity-PRS only captures a fraction of the variance in human longevity. However, the convergence of findings from the two approaches gives us confidence in the findings. Second, our replication sample, while well characterised, was relatively small, and some of the non-replicated findings were possibly due to low statistical power. The findings therefore should be replicated in independent larger samples. Third, we used a limited number of brain metrics. Several other brain volumetric measures as well as measures of brain microstructure (e.g. fractional anisotropy, mean diffusivity) are worthy of analysis in future work. Fourth, we did not examine intermediate phenotypes that link genetics with lifespan, several of which are disease related phenotypes that are important causes of death or infirmity. Finally, the generalisability of our results may not extend to other racial/ethnic groups, as we restricted our analyses to British ancestry.

In conclusion, lower WMH volumes were not only significantly associated with longer parental lifespan but were also associated with higher longevity-PRS, with stronger effects shown in the older-male group. Moreover, these effects were not driven exclusively by contributions from the *APOE* locus. The associations were not seen for cortical and grey matter volumes or hippocampal volumes. WMH may therefore be regarded as a marker of ageing and longevity. With WMH being markers of cerebrovascular disease, neurodegeneration and inflammation, this association is understandable, but the observed associations of WMH with longevity open up new possibilities for the better understanding of the pathways to longevity and healthy brain ageing.

## Data Availability

Raw data, including scans:
1. UK Biobank has made their data available to approved researchers and our data were obtained under approved application 37103.
2. Data of Sydney Memory and Ageing Study (MAS) and Older Australian Twins Study (OATS) will be made available to approved researchers via application to
Derived data, e.g. imaging-derived phonotypes (IDPs) will be made available after the publication of the manuscript by writing to the corresponding author
Computer programs/codes will be made available after the publication of the manuscript by writing to the corresponding author

## Acknowledgements

We would like to thank UK Biobank, MAS and OATS participants and the research teams of these projects. Also, we thank Angie Russell for her assistance in the preparation of the manuscript. This discovery research has been conducted using the UK Biobank resource (application number 37103). Ethics approval for the UK Biobank study was obtained from the North West Centre for Research Ethics Committee (11/NW/0382). Memory and Ageing Study (MAS) was supported by the National Health and Medical Research Council (NHMRC) Program Grant ID 350833. The Older Australian Twins Study (OATS) was supported by an NHMRC Strategic Award Grant of the Ageing Well, Ageing Productively Program (ID No. 401162). We gratefully acknowledge all the studies and databases that made their GWAS summary data available. Chao Dong is supported by the UNSW Scientia PhD Scholarship Program.

This research was undertaken with the assistance of resources and services from the National Computational Infrastructure (NCI), which is supported by the Australian Government.

