## Supplementary Materials for "Parental lifespan and polygenic risk score of longevity are associated with white matter hyperintensities"

Table S1. Scanning parameters in OATS.

| Scan site | Manufacturer | Scanner | Acquisition matrix size ($mm^3$) | Voxel size ($mm^3$) | TR (ms) | TE (ms) | TI (ms) | FA (degree) |
|---|---|---|---|---|---|---|---|---|
| **T1-weighted MRI** | | | | | | | | |
| New South Wales | Philips – 1.5T | Scanner 1 | 256×256×190 | 1×1×1 | 5.7 | 2.6 | 780 | 8 |
| New South Wales | Philips - 3T | Scanner 2 | 256×190×256 | 1×1×1 | 5.8 | 2.6 | 780 | 8 |
| Victoria | Siemens - 1.5T | Scanner 3 | 176×216×256 | 1×1.5×1 | 1530 | 3.2 | 780 | 8 |
| Queensland | Siemens - 3T | Scanner 4 | 208×240×256 | 1×1×1 | 2300 | 3.0 | 900 | 9 |
| Victoria | Siemens - 3T | Scanner 5 | 208×240×256 | 1×1×1 | 2300 | 3.0 | 900 | 9 |
| **T2-weighted FLAIR** | | | | | | | | |
| New South Wales | Philips – 1.5T | Scanner 1 | 256×256×133 | 0.5×0.5×3.5 | 11000 | 110 | 2800 | 90 |
| New South Wales | Philips - 3T | Scanner 2 | 198×256×256 | 0.6×1×1 | 4800 | 278.9 | 1650 | 90 |
| Victoria | Siemens - 1.5T | Scanner 3 | 184×230×150 | 0.9×0.9×3 | 10000 | 99 | 2500 | 120 |
| Queensland | Siemens - 3T | Scanner 4 | 182×224×165 | 0.7×0.7×3.3 | 9000 | 87 | 2500 | 150 |
| Victoria | Siemens - 3T | Scanner 5 | 208×256×132 | 0.4×0.4×3.3 | 9000 | 84 | 2500 | 150 |

Notes. TR: repetition time; TE: echo time; TI: inversion time; FA: flip angle.

Table S2. Maternal and paternal lifespan associations with brain metrics volumes.

| **Cohorts** | **Phenotypes** | **Maternal lifespan** | | | | **Paternal lifespan** | | | |
|---|---|---|---|---|---|---|---|---|---|
| | | β | SE | P | $P_{adj}$ | β | SE | P | $P_{adj}$ |
| **UK Biobank** (discovery sample) | Cortex | **0.0131** | **0.0047** | **0.0053*** | **0.0212*** | 0.0072 | 0.0042 | 0.0864 | 0.1383 |
| | Total Gray matter | **0.0139** | **0.0044** | **0.0017** | **0.0137*** | **0.0090** | **0.0040** | **0.0233*** | **0.0465*** |
| | Brain soft tissue | 0.0057 | 0.0038 | 0.1356 | 0.1809 | 0.0051 | 0.0034 | 0.1335 | 0.1780 |
| | Hippocampus | 0.0094 | 0.0072 | 0.1888 | 0.2157 | 0.0002 | 0.0064 | 0.9701 | 0.9849 |
| | Cerebral WM | -0.0036 | 0.0047 | 0.4412 | 0.4412 | 0.0001 | 0.0043 | 0.9849 | 0.9849 |
| | Whole WMH | -0.0171 | 0.0087 | 0.0511 | 0.1022 | **-0.0347** | **0.0078** | **8.51E-06*** | **0.0001*** |
| | PVWMH | -0.0151 | 0.0088 | 0.0850 | 0.1359 | **-0.0330** | **0.0078** | **2.50E-05*** | **0.0001*** |
| | DWMH | -0.0187 | 0.0090 | 0.0379* | 0.1011 | **-0.0289** | **0.0080** | **0.0003*** | **0.0009*** |
| **MAS & OATS** (replication sample) | Cortex | -0.0052 | 0.0281 | 0.8541 | 0.8541 | 0.0343 | 0.0282 | 0.2248 | 0.4939 |
| | Total Gray matter | -0.0068 | 0.0260 | 0.7949 | 0.8541 | 0.0264 | 0.0261 | 0.3124 | 0.4939 |
| | Brain soft tissue | -0.0126 | 0.0229 | 0.5823 | 0.8541 | 0.0326 | 0.0229 | 0.1559 | 0.4939 |
| | Hippocampus | 0.0130 | 0.0322 | 0.6872 | 0.8541 | -0.0248 | 0.0322 | 0.4411 | 0.4939 |
| | Cerebral WM | -0.0075 | 0.0233 | 0.7468 | 0.8541 | 0.0338 | 0.0233 | 0.1466 | 0.4939 |
| | Whole WMH | -0.0775 | 0.0322 | 0.0164* | 0.0654 | -0.0279 | 0.0322 | 0.3858 | 0.4939 |
| | PVWMH | -0.0856 | 0.0322 | 0.0081* | 0.0645 | -0.0220 | 0.0322 | 0.4939 | 0.4939 |
| | DWMH | -0.0478 | 0.0342 | 0.1623 | 0.4329 | -0.0365 | 0.0341 | 0.2852 | 0.4939 |

Notes. Brain soft tissue (labelled as BrainSegVolNotVent in FreeSurfer); WM: white matter; WMH: white matter hyperintensity; PVWMH: periventricular WMH; DWMH: deep WMH. Asterisks represent statistical significance when using linear mixed models ($P < 0.05$). SE: standard error. $P_{adj}$: corrected for False Discovery Rate. MAS: Sydney Memory and Ageing Study; OATS: Older Australian Twins Study.

Table S3. Quartile analyses.

| Cohorts | Phenotypes | Quartiles: lowest PRS group *VS* highest PRS group | | | |
|---|---|---|---|---|---|
| | | β | SE | P | $P_{adj}$ |
| **UK Biobank** (discovery sample) | Whole WMH | **0.0571** | **0.0204** | **0.0052*** | **0.0157*** |
| | PVWMH | **0.0436** | **0.0206** | **0.0342*** | **0.0494*** |
| | DWMH | **0.0761** | **0.0211** | **0.0003*** | **0.0056*** |
| | Whole WMH (no *APOE*) | **0.0452** | **0.0205** | **0.0275*** | **0.0494*** |
| | PVWMH (no *APOE*) | 0.0330 | 0.0207 | 0.1106 | 0.1106 |
| | DWMH (no *APOE*) | **0.0640** | **0.0212** | **0.0025*** | **0.0157*** |

Notes. WMH: white matter hyperintensity; PVWMH: periventricular WMH; DWMH: deep WMH. Asterisks represent statistical significance when using linear mixed models ($P < 0.05$). SE: standard error. $P_{adj}$: corrected for False Discovery Rate.

Table S4. Associations between brain metrics volumes and interaction of age and sex (age × sex) in the longevity-PRS analysis.

| **Phenotypes** | **UK Biobank** (discovery sample) | | | | | | | |
|---|---|---|---|---|---|---|---|---|
| | β (with/no *APOE*) | | SE (with/no *APOE*) | | P (with/no *APOE*) | | $P_{adj}$ (with/no *APOE*) | |
| Cortex | **-0.0901** | **-0.0901** | **0.0077** | **0.0077** | **2.28E-31*** | **2.29E-31*** | **6.11E-31*** | **6.11E-31*** |
| Total grey matter | **-0.0972** | **-0.0973** | **0.0072** | **0.0072** | **5.89E-41*** | **5.77E-41*** | **2.36E-40*** | **2.36E-40*** |
| Brain soft tissue | **-0.0971** | **-0.0971** | **0.0062** | **0.0062** | **1.46E-54*** | **1.45E-54*** | **1.17E-53*** | **1.17E-53*** |
| Hippocampus | **-0.0906** | **-0.0869** | **0.0118** | **0.0078** | **1.95E-14*** | **6.93E-29*** | **3.14E-14*** | **1.39E-28*** |
| Cerebral WM | **-0.069** | **-0.0905** | **0.0078** | **0.0118** | **6.94E-29*** | **1.96E-14*** | **1.39E-28*** | **3.14E-14*** |
| Whole WMH | **-0.0600** | **-0.0602** | **0.0142** | **0.0142** | **2.27E-05*** | **2.16E-05*** | **3.03E-05*** | **3.03E-05*** |
| PVWMH | **-0.0581** | **-0.0583** | **0.0143** | **0.0143** | **4.65E-05*** | **4.47E-05*** | **5.31E-05*** | **5.31E-05*** |
| DWMH | **-0.0591** | **-0.0592** | **0.0146** | **0.0146** | **5.31E-05*** | **5.05E-05*** | **5.31E-05*** | **5.31E-05*** |

Notes. Values of β, SE, P, $P_{adj}$ in the analysis with and no *APOE* were presented in the table. Brain soft tissue (labelled as BrainSegVolNotVent in FreeSurfer); WM: white matter; WMH: white matter hyperintensity; PVWMH: periventricular WMH, DWMH: deep WMH. Asterisks represent statistical significance when using linear mixed models ($P < 0.05$). SE: standard error. $P_{adj}$: corrected for False Discovery Rate.

Table S5. Age and sex differences in associations between brain volumes and longevity-PRS in the UK Biobank sample.

| **Cohorts** | **Phenotypes** | **Male** (with/no *APOE*) | | | | | | | | **Female** (with/no *APOE*) | | | | | | | |
|---|---|---|---|---|---|---|---|---|---|---|---|---|---|---|---|---|---|
| | | β | | SE | | P | | $P_{adj}$ | | β | | SE | | P | | $P_{adj}$ | |
| **All UK Biobank participants** | Cortex | **0.0202** | **0.0233** | **0.0086** | **0.0086** | **0.0183*** | **0.0066*** | 0.0915 | 0.0656 | -0.0105 | -0.0095 | 0.0075 | 0.0075 | 0.1613 | 0.2066 | 0.6438 | 0.6438 |
| | Total Gray matter | 0.0108 | 0.0136 | 0.0082 | 0.0083 | 0.1916 | 0.1004 | 0.3624 | 0.3346 | -0.0097 | -0.0082 | 0.0072 | 0.0072 | 0.1772 | 0.2575 | 0.6438 | 0.6438 |
| | Brain soft tissue | 0.0088 | 0.0103 | 0.0072 | 0.0072 | 0.2174 | 0.1526 | 0.3624 | 0.3624 | -0.0045 | -0.0028 | 0.0062 | 0.0062 | 0.4675 | 0.6474 | 0.9351 | 0.9425 |
| | Hippocampus | 0.0050 | 0.0064 | 0.0120 | 0.0120 | 0.6787 | 0.5941 | 0.6787 | 0.6601 | 0.0034 | 0.0048 | 0.0109 | 0.0109 | 0.7547 | 0.6597 | 0.9425 | 0.9425 |
| | Cerebral WM | 0.0056 | 0.0060 | 0.0087 | 0.0087 | 0.5155 | 0.4893 | 0.6444 | 0.6444 | -0.0003 | 0.0015 | 0.0077 | 0.0077 | 0.9663 | 0.8483 | 0.9663 | 0.9425 |
| **Over 63 age group in UK Biobank** | Cortex | 0.0135 | 0.0160 | 0.0101 | 0.0101 | 0.1789 | 0.1124 | 0.6773 | 0.6773 | -0.0013 | 0.0003 | 0.0102 | 0.0102 | 0.8980 | 0.9796 | 0.9796 | 0.9796 |
| | Total Gray matter | 0.0040 | 0.0064 | 0.0097 | 0.0097 | 0.6773 | 0.5094 | 0.6773 | 0.6773 | 0.0015 | 0.0018 | 0.0097 | 0.0097 | 0.8798 | 0.8523 | 0.9796 | 0.9796 |
| | Brain soft tissue | 0.0050 | 0.0070 | 0.0084 | 0.0084 | 0.5531 | 0.4054 | 0.6773 | 0.6773 | 0.0052 | 0.0054 | 0.0083 | 0.0083 | 0.5335 | 0.5158 | 0.9796 | 0.9796 |
| | Hippocampus | 0.0088 | 0.0120 | 0.0141 | 0.0142 | 0.5354 | 0.3969 | 0.6773 | 0.6773 | 0.0169 | 0.0175 | 0.0143 | 0.0143 | 0.2371 | 0.2210 | 0.9796 | 0.9796 |
| | Cerebral WM | 0.0050 | 0.0068 | 0.0101 | 0.0101 | 0.6205 | 0.5023 | 0.6773 | 0.6773 | 0.0045 | 0.0046 | 0.0103 | 0.0103 | 0.6626 | 0.6513 | 0.9796 | 0.9796 |
| **Under 63 age group in UK Biobank** | Cortex | **0.0319** | **0.0366** | **0.0159** | **0.0159** | **0.0443*** | **0.0214*** | 0.2215 | 0.2141 | -0.0207 | -0.0203 | 0.0115 | 0.0116 | 0.0728 | 0.0794 | 0.2213 | 0.2213 |
| | Total Gray matter | 0.0234 | 0.0271 | 0.0153 | 0.0153 | 0.1264 | 0.0770 | 0.3160 | 0.2566 | -0.0219 | -0.0189 | 0.0111 | 0.0111 | 0.0478* | 0.0885 | 0.2213 | 0.2213 |
| | Brain soft tissue | 0.0157 | 0.0161 | 0.0131 | 0.0132 | 0.2308 | 0.2205 | 0.3846 | 0.3846 | -0.0148 | -0.0115 | 0.0095 | 0.0096 | 0.1192 | 0.2305 | 0.2384 | 0.3841 |
| | Hippocampus | -0.0031 | -0.0051 | 0.0225 | 0.0226 | 0.8916 | 0.8228 | 0.8916 | 0.8916 | -0.0119 | -0.0095 | 0.0172 | 0.0173 | 0.4898 | 0.5813 | 0.6678 | 0.6678 |
| | Cerebral WM | 0.0072 | 0.0049 | 0.0154 | 0.0155 | 0.6417 | 0.7529 | 0.8916 | 0.8916 | -0.0061 | -0.0024 | 0.0117 | 0.0117 | 0.6010 | 0.8404 | 0.6678 | 0.8404 |

Notes. Values of β, SE, P, $P_{adj}$ in the analysis with and no *APOE* were presented in the table. Brain soft tissue (labelled as BrainSegVolNotVent in FreeSurfer). Asterisks represent statistical significance when using linear mixed models ($P < 0.05$). SE: standard error. $P_{adj}$: corrected for False Discovery Rate. MAS: Sydney Memory and Ageing Study; OATS: Older Australian Twins Study.

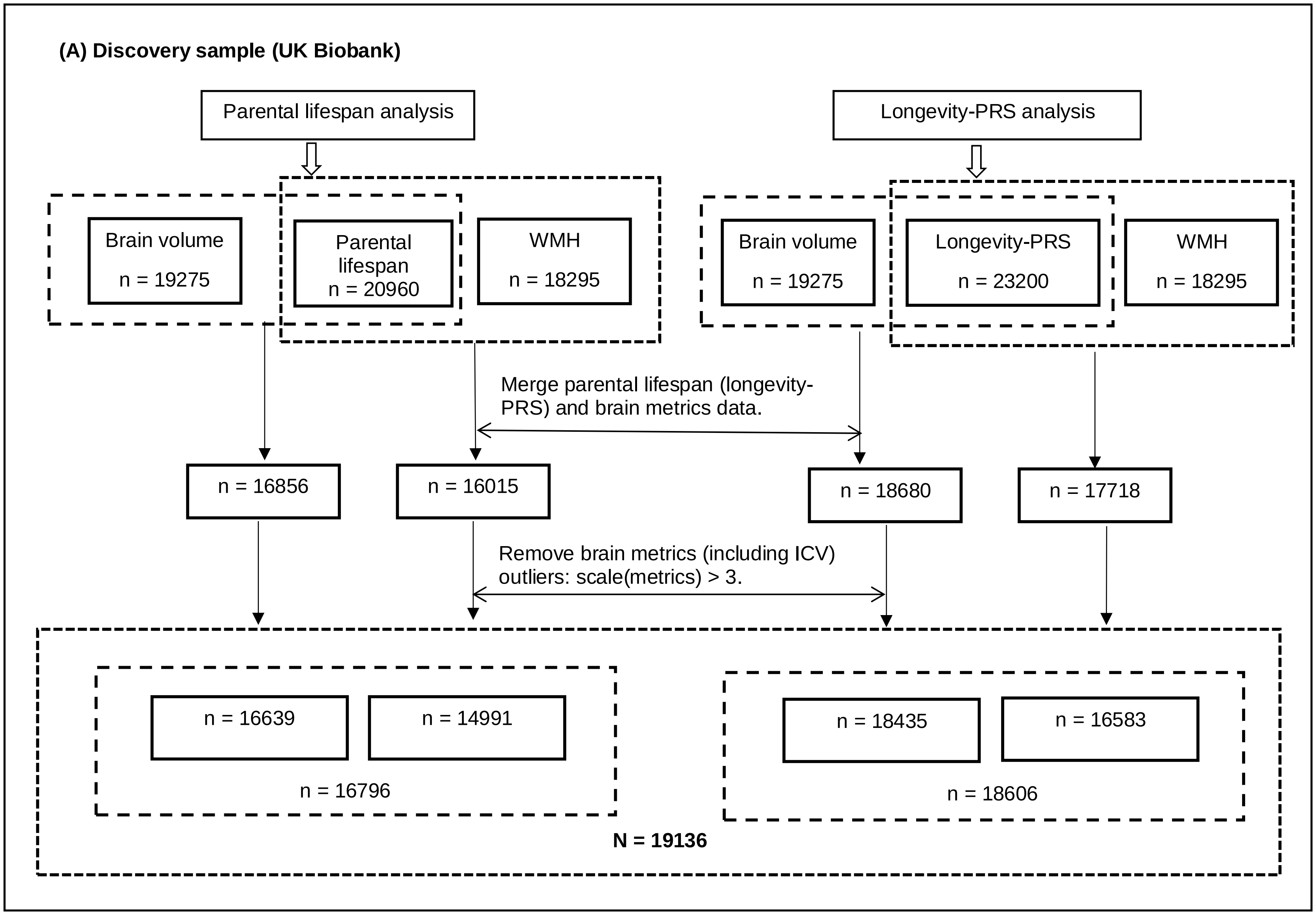
(A) Discovery sample (UK Biobank)
Parental lifespan analysis
Longevity-PRS analysis
Brain volume n = 19275
Parental lifespan n = 20960
WMH n = 18295
Brain volume n = 19275
Longevity-PRS n = 23200
WMH n = 18295
Merge parental lifespan (longevity-PRS) and brain metrics data.
n = 16856
n = 16015
n = 18680
n = 17718
Remove brain metrics (including ICV) outliers: scale(metrics) > 3.
n = 16639
n = 14991
n = 16796
n = 18435
n = 16583
n = 18606
N = 19136

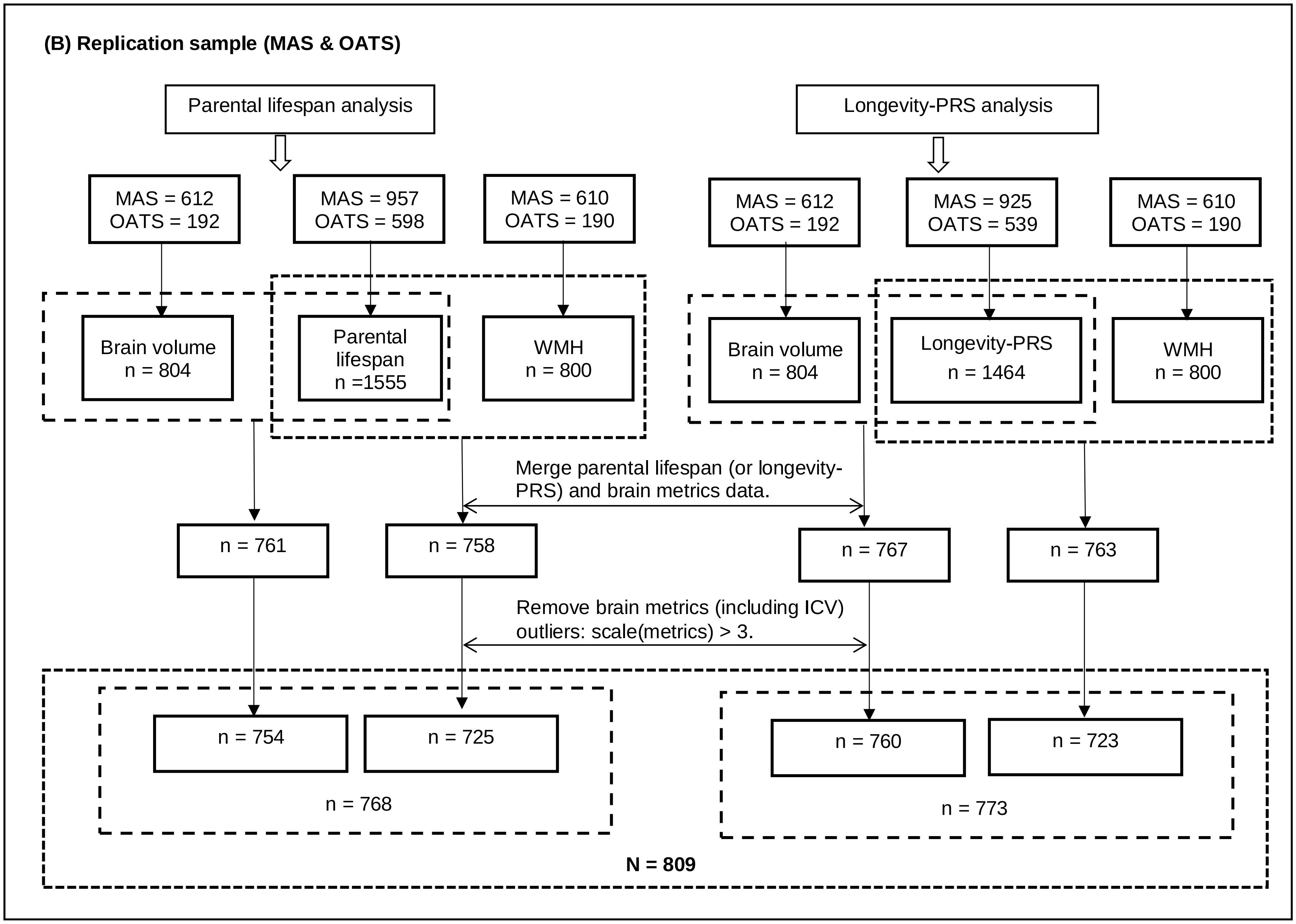


**Figure S1**. **Analytic sample derivation of discovery sample and replication sample**. In each dataset, there were two analyses undertaken: parental lifespan analysis and longevity-PRS analysis. In each analysis, association tests were performed with brain volumes and WMH. (A) Discovery sample. In the parental lifespan analysis, only participants having their parental age at death data and brain volume (or WMH) data were included. After excluding brain metrics outliers (scale (metrics) > 3), the final analysis included 16796 participants. Similarly, in the longevity-PRS analysis, only participants having longevity-PRS data and brain volume (or WMH) data were included. After excluding brain metrics outliers (scale (metrics) > 3), the final analysis included 18606 participants. (B) Replication sample. Using the same procedure as used in the discovery sample, the final analysis included 768 participants in the parental lifespan analysis and 773 participants in the longevity-PRS analysis.

MAS: Sydney Memory and Ageing Study; OATS: Older Australian Twins Study.
